## Supplementary Information for "Risk assessment of COVID-19 epidemic resurgence in relation to SARS-CoV-2 variants and vaccination passes"

### Risk assessment of COVID-19 epidemic resurgence in relation to SARS-CoV-2 variants and vaccination passes Supplementary Text, Tables and Figures

Tyll Krueger[1,†], Krzysztof Gogolewski[2,†], Marcin Bodych[1,†], Anna Gambin[2], Giulia Giordano[3], Sarah Cuschieri[4], Thomas Czypionka[5,6], Matjaz Perc[7,8,9,10], Elena Petelos[11,12], Magdalena Rosińska[13], and Ewa Szczurek[2,*]

[1]*Faculty of Electronics, Department of Control Systems and Mechatronics, Wroclaw University of Science and Technology, Wroclaw, Poland*

[2]*Faculty of Mathematics, Informatics and Mechanics, University of Warsaw, Warsaw, Poland*

[3]*Department of Industrial Engineering, University of Trento, Trento, Italy*

[4]*Department of Anatomy, Faculty of Medicine and Surgery, University of Malta, Msida, Malta*

[5]*Institute for Advanced Studies, Josefstädterstraße 39, 1080, Vienna, Austria*

[6]*London School of Economics and Political Science, Houghton Street, WC2A 2AE, London, UK.*

[7]*Faculty of Natural Sciences and Mathematics, University of Maribor, Koroška cesta 160, 2000 Maribor, Slovenia,* [8]*Complexity Science Hub Vienna, Josefstädterstraße 39, 1080 Vienna, Austria,* [9]*Department of Medical Research, China Medical University Hospital, China Medical University, Taichung 404332, Taiwan,* [10]*Alma Mater Europaea, Slovenska ulica 17, 2000 Maribor, Slovenia*

[11]*Clinic of Social and Family Medicine, Faculty of Medicine, University of Crete, Heraklion, Greece,* [12]*Department of Health Services Research, CAPHRI-Care and Public Health Research Institute, Maastricht University, Maastricht, The Netherlands*

[13]*Department of Infectious Disease Epidemiology and Surveillance, National Institute of Public Health, Warsaw, Poland.*

#### Supplementary Note 1

##### Endemic state

The endemic state of the VAP-SIRS model is obtained as the solution of the system of equations defined by setting the derivatives of the ODE system (1) to 0. A straightforward computation reduces the endemic system of equations to the following:

$$
\begin{aligned}
0 &= \left(\beta I + \delta^{+} I_V\right)\left(d - I - \frac{\gamma}{\kappa} I\right) - \gamma I && (1)\\
0 &= \frac{\upsilon_r \alpha \left(\omega + \beta I + \delta^{*} I_V\right) \cdot \gamma I_V}{\left(\beta I + \delta^{*} I_V + \omega + \upsilon_r \left(1 - \alpha\right)\right)} \\
&\quad - \omega \left(1 - d - I_V - \frac{\gamma}{\kappa + \upsilon_r} I_V\right)\left(\beta I + \delta^{*} I_V\right) \\
&\quad + \omega \gamma I_V + \upsilon_r \frac{\gamma}{\kappa + \upsilon_r} I_V \cdot \left(\beta I + \delta^{*} I_V\right) && (2)\\
S &= d - I\left(1 + \frac{\gamma}{\kappa}\right) && (3)\\
S_V &= \frac{\gamma I_V}{\beta I + \delta^{*} I_V} && (4)\\
R &= \frac{\gamma}{\kappa} I && (5)\\
R_V &= \frac{\gamma}{\kappa + \upsilon} I_V && (6)\\
V &= 1 - d - S - S_V - I - I_V - R - R_V, && (7)
\end{aligned}
$$

where the variables $\delta^{*}, \delta^{+}$ are defined separately for each mixing type. For the proportional mixing:

$$\delta^{*} = \delta \quad \text{and} \quad \delta^{+} = \beta, \tag{8}$$

while for the preferential mixing:

$$\delta^{*} = \frac{\delta^2}{\beta d + \delta\left(1 - d\right)} \quad \text{and} \quad \delta^{+} = \frac{\beta^2}{\beta d + \delta\left(1 - d\right)}. \tag{9}$$

**Hospitalizations in the endemic state**

We consider a simplified model for hospitalizations, where we introduce the hospitalized compartments $H_1$, $H_2$ and $H_D$ as subgroups of the infected compartments $I_1$, $I_2$, and $I_D$, respectively. Similarly, we introduce the subgroups $S_{1H}$, $S_{2H}$ and $S_{DH}$ of susceptibles that lack protection against hospitalization. The mean duration of being infectious is the same for all hospitalized and the remaining individuals in the infected compartments. Hence, the additional compartmentalization does not affect the overall dynamics of the ODE system (1) described in the main text. Note that, in the endemic state, $S_N = R_N = I_N = 0$.

Denote the waning rate of protection against severe progression as $\omega_H$. Recent findings from Public Health England [1], suggest that, for the vaccinated population, a 10% loss of protection against hospitalization occurs within six months post-vaccination. We thus set $\omega_H = 0.001$.

Let $\alpha_H$ be the conditional effectiveness of protection against hospitalization given that an individual has lost its protection against infection, or has never built up protection against infection after vaccination. To estimate $\alpha_H$, we use the German data available as of 02.09.2021, reported by the Robert Koch Institute [2]. Specifically, we consider values: $a \doteq 1-\alpha$, the *a-priori* likelihood for a vaccinated to get infected if exposed to an infectious contact, and $b \doteq (1-\alpha_H)(1-\alpha)$, the *a-priori* likelihood to get infected and hospitalized in the event of an infectious contact [3]. These data and the corresponding $\alpha_H$ values can be organized in the following table, specified for two age groups: people aged 60 years and older and group of adults aged 18 to 59 years.

$$\begin{pmatrix} age & a & b & \alpha_H \\ \geq 60 & 0.17 & 0.06 & 0.65 \\ 18-59 & 0.16 & 0.05 & 0.69 \end{pmatrix}. \tag{10}$$

In view of the above values extracted from real-world data, we assume a conservative value of $\alpha_H = 0.7$ in the simulations. The time duration of hospitalization is assumed to be exponentially distributed with mean denoted by $1/\tau_H$. Given recent reports [4] we assume $1/\tau_H = 17$.

Finally, we introduce $\theta$ as the likelihood for an unprotected susceptible to be hospitalized in case of infection. $\theta$ is set to 0.03 based on recent reports on hospitalization rates among reported cases [5], and under the assumption that the total (including unreported) number of cases is larger by a factor of two.

Due to lack of data needed to estimate variant-specific values, the four parameters discussed above ($\omega_H$, $\alpha_H$, $1/\tau_H$, and $\theta$) are set to the same values for both the Alpha and the Delta variants.

We have the following set of equations:

$$0 = \frac{dH_1}{dt} = \theta(\beta I + \beta_V I_V) S_{1H} - \tau_H H_1 \tag{11}$$

$$0 = \frac{dH_2}{dt} = \theta(\beta I + \beta_V I_V) S_{2H} - \tau_H H_2 \tag{12}$$

$$0 = \frac{dH_D}{dt} = \theta(\beta I + \beta_V I_V) S_{DH} - \tau_H H_D \tag{13}$$

$$0 = \frac{dS_{DH}}{dt} = -(\beta I + \beta_V I_V) S_{DH} + \omega_H (S_D - S_{DH}) + (1-\alpha_H)\kappa R_D \tag{14}$$

$$0 = \frac{dS_{1H}}{dt} = -(\beta I + \beta_V I_V) S_{1H} + \omega_H (S_1 - S_{1H}) + \upsilon_r (1-\alpha_H)(1-\alpha) S_{2H} - \omega S_{1H} \tag{15}$$

$$0 = \frac{dS_{2H}}{dt} = -(\beta I + \beta_V I_V) S_{2H} + (1-\alpha_H)\omega V + \omega_H (S_2 - S_{1H}) + \tag{16}$$

$$+\omega S_{1H} + (1-\alpha_H)\kappa R_V - \upsilon_r S_{2H}. \tag{17}$$

The explicit solution for $S_{DH}, S_{1H}, S_{2H}$ is straightforward and we have

$$H_1 + H_2 + H_D = \frac{1}{\tau_H}\theta\left(\beta I + \beta_V I_V\right)\left(S_{1H} + S_{2H} + S_{DH}\right). \qquad (18)$$

Assuming preferential mixing, instead of proportional mixing, leads to the same changes as in the basic ODE model, namely, the following replacements have to be made:

$$S_{1H}I_V \quad \text{is replaced by} \quad \frac{\beta_v}{\beta\left(S+I+R\right)+\beta_v\left(1-\left(S+I+R\right)\right)}S_{1H}I_V \qquad (19)$$

$$S_{2H}I_V \quad \text{is replaced by} \quad \frac{\beta_v}{\beta\left(S+I+R\right)+\beta_v\left(1-\left(S+I+R\right)\right)}S_{2H}I_V \qquad (20)$$

$$S_{DH}I_V \quad \text{is replaced by} \quad \frac{\beta}{\beta\left(S+I+R\right)+\beta_v\left(1-\left(S+I+R\right)\right)}S_{DH}I_V. \qquad (21)$$

#### Supplementary Tables

| | Parameter setup | $a$ | $v_r$ | $d$ | $\omega$ | $V^{\mathrm{as}}$ | $f_{\min}$ |
|---|---|---|---|---|---|---|---|
| 1 | Reference setup (ref Fig. S2a) | 0.92 | 0.004 | 0.12 | 0.002 | 0.54 | 0.46 |
| 2 | Decreased $a$ (ref Fig. S2b) | **0.73** | 0.004 | 0.12 | 0.002 | 0.43 | 0.56 |
| 3 | Increased $v_r$ (ref Fig. S2c) | 0.92 | **0.008** | 0.12 | 0.002 | 0.65 | 0.29 |
| 4 | Increased $d$ (ref Fig. S2e) | 0.92 | 0.004 | **0.3** | 0.002 | 0.43 | 0.56 |
| 5 | Increased $\omega$ (ref Fig. S2f) | 0.92 | 0.004 | 0.12 | **0.005** | 0.36 | 0.61 |
| 6 | Decreased $a$ and increased $v_r$ (ref Fig. S4a) | **0.73** | **0.008** | 0.12 | 0.002 | 0.51 | 0.49 |
| 7 | Decreased $a$ and increased $d$ (ref Fig. S4b) | **0.73** | 0.004 | **0.3** | 0.002 | 0.34 | 0.62 |
| 8 | Decreased $a$ and increased $\omega$ (ref Fig. S4c) | **0.73** | 0.004 | 0.12 | **0.005** | 0.29 | 0.65 |
| 9 | Increased $v_r$, increased $d$ (ref Fig., S4d) | 0.92 | **0.008** | **0.3** | 0.002 | 0.52 | 0.48 |
| 10 | Increased $v_r$, increased $\omega$ (ref Fig., S4e) | 0.92 | **0.008** | 0.12 | **0.005** | 0.50 | 0.50 |
| 11 | Increased $d$, increased $\omega$ (ref Fig., S4f) | 0.92 | 0.004 | **0.3** | **0.005** | 0.21 | 0.68 |

Supplementary Table S1: **Asymptotic level of immunization $V^{\mathrm{as}}$ and minimum common restrictions $f_{\min}$ for the Alpha variant different parameter setups**, for parameters: vaccine effectiveness $a$, revaccination rate $v_r$, fraction of never-vaccinated $d$, and waning immunity rate $\omega$. The first row concerns the reference setup for the Alpha variant; rows below are setups with the same parameters as in the reference setup, but with either one parameter changed (in bold; rows 2–5; same as in Fig. S2, apart from preferential mixing, as it is not relevant for common restrictions) or two parameters changed (in bold; rows 6–11; same as in Fig. S4).

#### Supplementary Figures

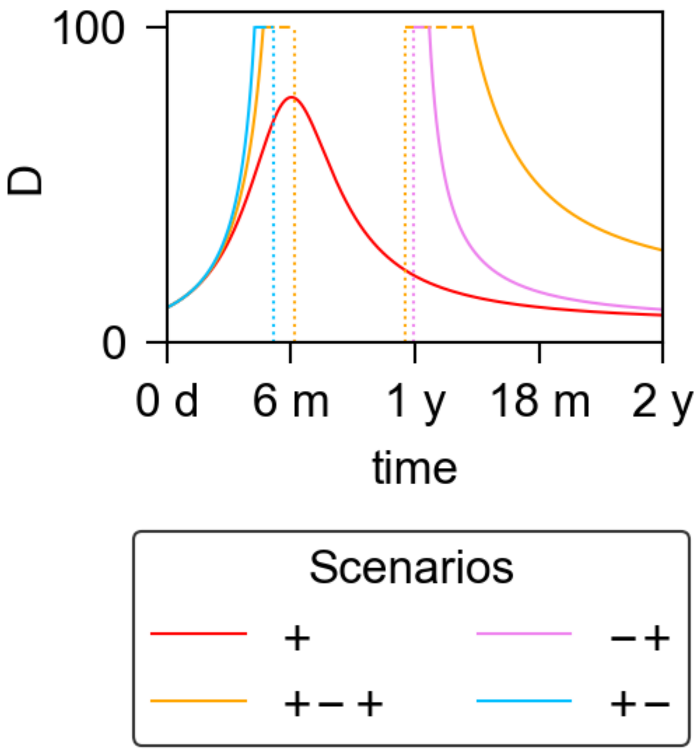


Supplementary Figure S1: **Instantaneous doubling times for the examples of five scenarios for the Delta variant, depending on the restrictions for VP holders and for the rest of the population.** For the reference setup for the Delta variant ($a = 0.79$ – corresponding to the effectiveness of the Comirnaty vaccine on the Delta variant, $\upsilon = \upsilon_r = 0.004$, $\omega = 0.002$, $d = 0.12$ – corresponding to the fraction of never-vaccinated in the United Kingdom, and proportional mixing), we compare the instantaneous doubling time $D$ (y axis, in days) as a function of time (x axis) for four different scenarios describing the epidemic evolution: overcritical (+, red, $f = 0.77$ and $f_{\text{v}} = 0.38$), initially and eventually overcritical (+−+, orange, $f = 0.77$ and $f_{\text{v}} = 0.55$), eventually overcritical (−+, pink, $f = 0.92$ and $f_{\text{v}} = 0.38$), and eventually subcritical (+−, cyan, with $f = 0.77$ and $f_{\text{v}} = 0.71$). The doubling curves are cut above 100 days, in the time intervals indicated by the horizontal dashed lines. Negative doubling times (halving times) are not plotted. Thus, the subcritical scenario (−) is not visualised. Vertical dotted lines indicate the times at which a transition occurs from overcritical to subcritical, or vice versa.

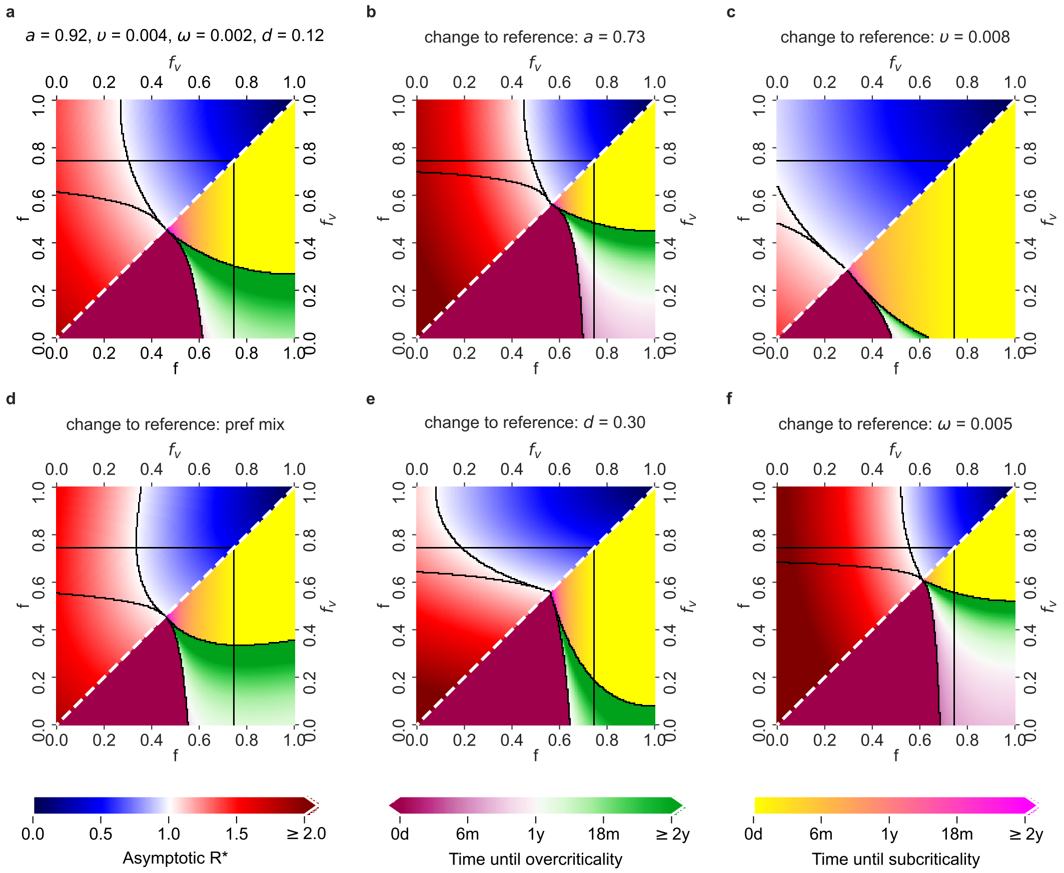


Supplementary Figure S2: **Possible COVID-19 epidemic dynamics for different parameter setups for the Alpha variant.** Lower triangles show the time until the last critical threshold, while upper triangles show the asymptotic $\mathcal{R}^*$, both as functions of the values of $f$ and $f_v$. Colors of the lower and upper triangles as in Figure 2 in the main text. **a.** Reference setup for the Alpha variant, with $a = 0.92$ (corresponding to the effectiveness of the Comirnaty vaccine on the Alpha variant), $\upsilon = \upsilon_r = 0.004$, $\omega = 0.002$, $d = 0.12$ (corresponding to the fraction of never-vaccinated in the United Kingdom), and proportional mixing. **b.** Setup with decreased vaccine effectiveness: $a = 0.73$ (the effectiveness of Vaxzevria on the Alpha variant). **c.** Setup with increased (re-)vaccination rate: $\upsilon = \upsilon_r = 0.008$. **d.** Setup with preferential (instead of proportional) mixing. **e.** Setup with increased fraction of people who will not get vaccinated: $d = 0.30$ (fraction of never-vaccinated in France). **f.** Setup with increased waning rate: $\omega = 0.005$.

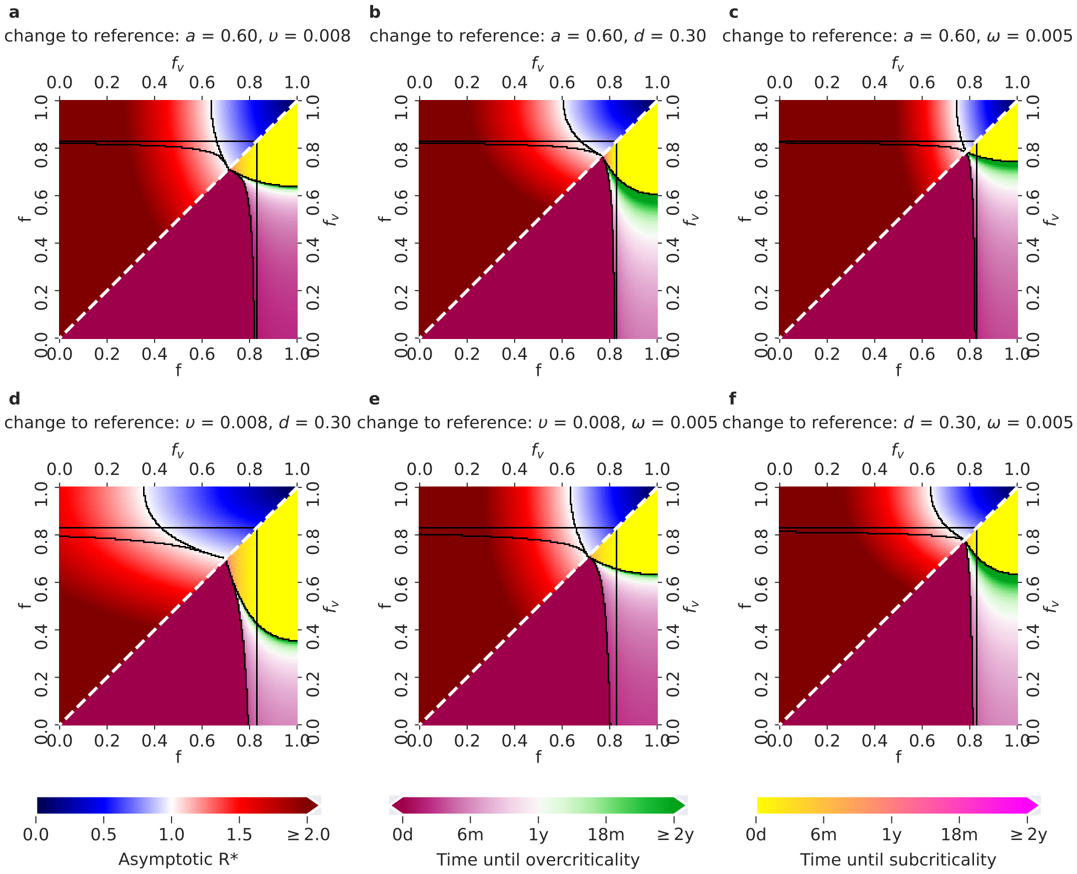


Supplementary Figure S3: **Possible COVID-19 epidemic dynamics for parameter setups with two changes w.r.t. the reference setup, for the Delta variant (changes as in main text Table 1, rows 6-11)** Colors of the lower triangles correspond to the time until critical changes in epidemic dynamics, while the colors of the upper triangles correspond to the values of asymptotic $\mathcal{R}^*$, as in Figure 2 in the main text. **a.** Setup with decreased effectiveness $a = 0.60$ (corresponding to the effectiveness of the Vaxzevira vaccine on the Delta variant), and increased (re-)vaccination rates $\upsilon = \upsilon_r = 0.008$. **b.** Setup with decreased $a = 0.60$ and increased fraction of never vaccinated $d = 0.30$ (corresponding to the change from the fraction of never-vaccinated in the United Kingdom to the fraction of never-vaccinated in France). **c.** Setup with decreased $a = 0.60$ and increased waning rate $\omega = 0.005$. **d.** Setup with increased $\upsilon = \upsilon_r = 0.008$ and increased $d = 0.30$. **e.** Setup with increased $\upsilon = \upsilon_r = 0.008$ and increased $\omega = 0.005$. **f.** Setup with increased $d = 0.30$ and increased $\omega = 0.005$.

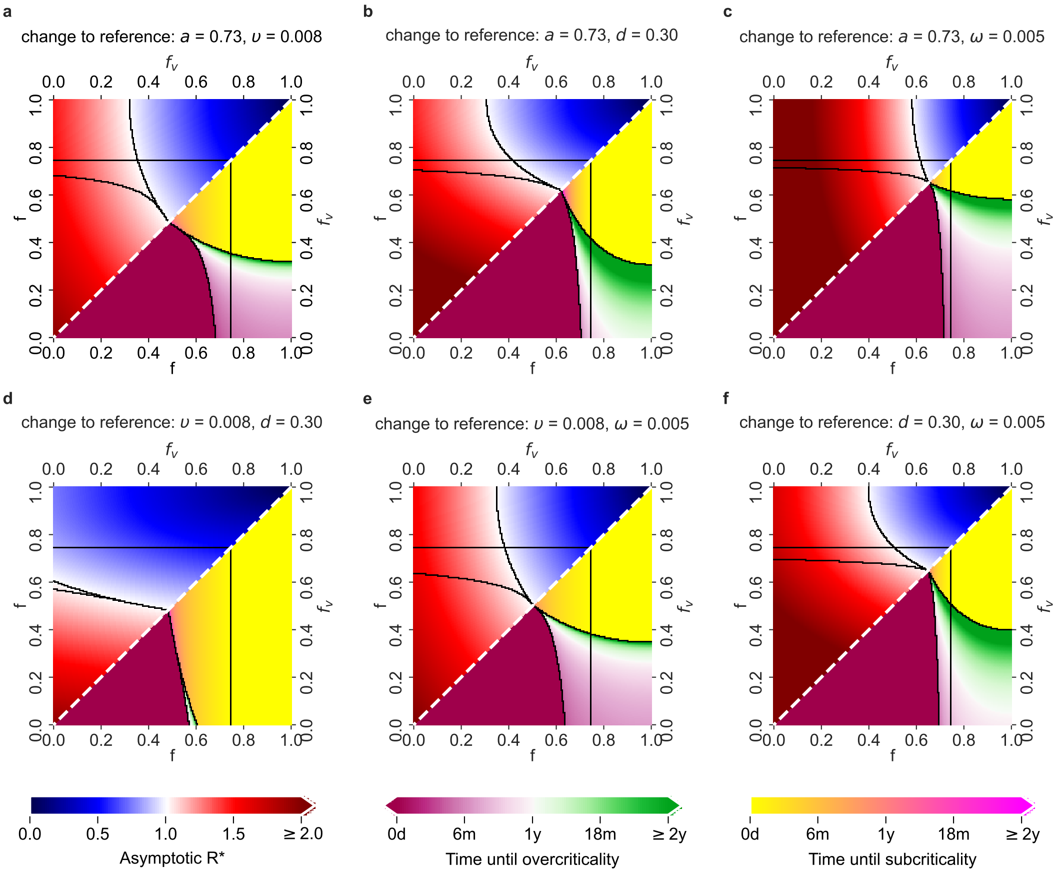


Supplementary Figure S4: **Possible COVID-19 epidemic dynamics for parameter setups with two changes w.r.t. the reference setup, for the Alpha variant.** Lower triangles show the time until the last critical threshold, while upper triangles show the asymptotic $\mathcal{R}^*$, both as functions of the values of $f$ and $f_v$. Colors of the lower and upper triangles as in Figure 2 in the main text. **a.** Setup with decreased effectiveness $a = 0.73$ (corresponding to the effectiveness of the Vaxzevira vaccine on the Alpha variant), and increased (re-)vaccination rates $\upsilon = \upsilon_r = 0.008$. **b.** Setup with decreased $a = 0.73$ and increased fraction of never vaccinated $d = 0.30$ (fraction of never-vaccinated in France). **c.** Setup with decreased $a = 0.73$ and increased waning rate $\omega = 0.005$. **d.** Setup with increased $\upsilon = \upsilon_r = 0.008$ and increased $d = 0.30$ **e.** Setup with increased $\upsilon = \upsilon_r = 0.008$ and increased $\omega = 0.005$. **f.** Setup with increased $d = 0.30$ and increased $\omega = 0.005$.

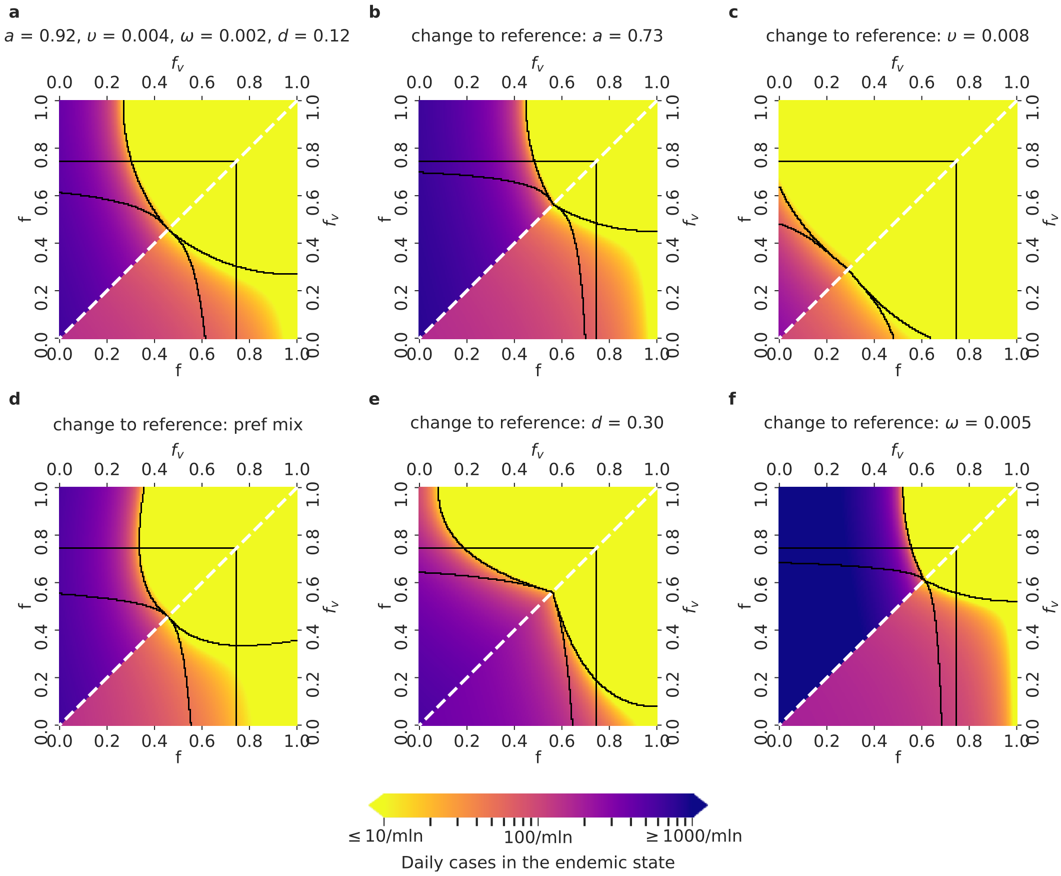


Supplementary Figure S5: **Daily COVID-19 infection cases in the endemic state, for the Alpha variant.** Lower triangles show the daily infection numbers in the unvaccinated, and upper triangles in the vaccinated population in the endemic state of the epidemics, for the relevant $f - f_{\text{v}}$ parameter space, where $f_{\text{v}} \leq f$ (as in Figure 4 in the main text). Parameter setups in panels **a.–f.** as in Supplementary Fig. S2.

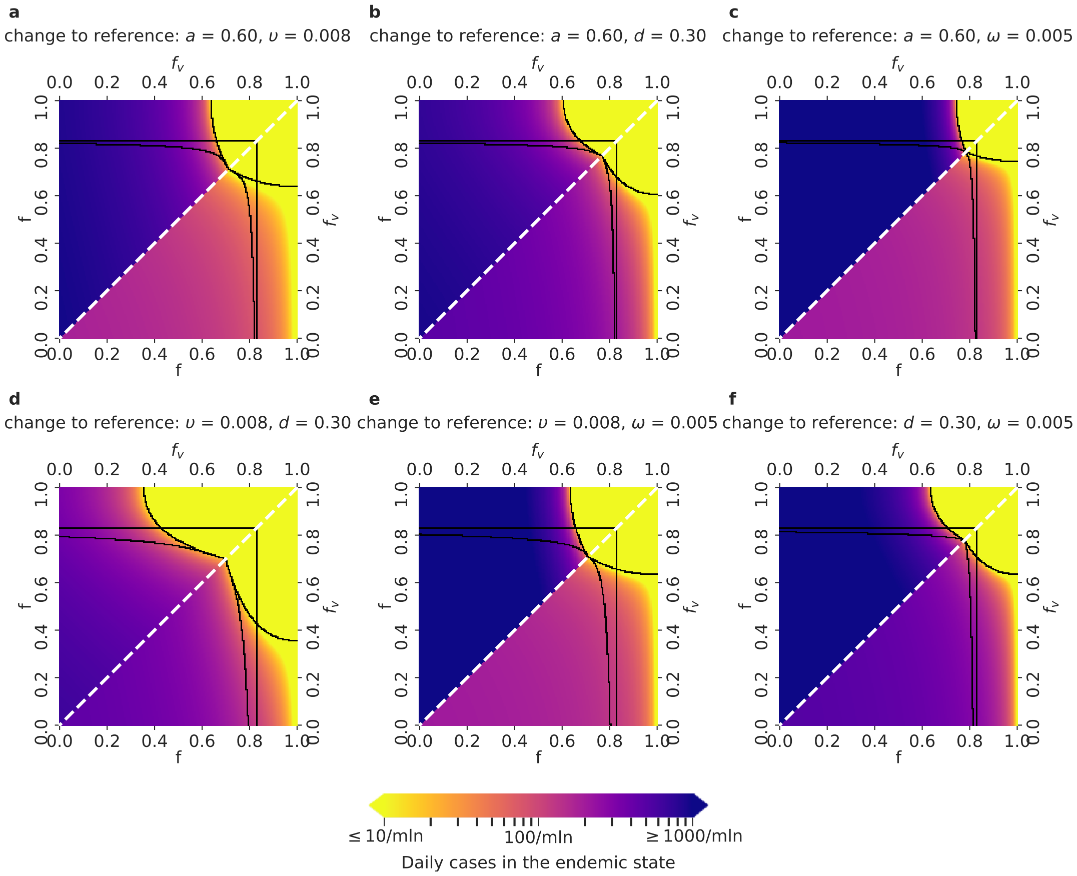


Supplementary Figure S6: **Daily COVID-19 infection cases in the endemic state for parameter setups with two changes w.r.t. the reference setup, and the Delta variant.** Lower triangles show the daily infection numbers in the unvaccinated, and upper triangles in the vaccinated population in the endemic state of the epidemics, for the relevant $f - f_{\mathrm{v}}$ parameter space, where $f_{\mathrm{v}} \leq f$. Black borders that delineate the five regions and parameter setups are identified as in the Supplementary Figure S3.

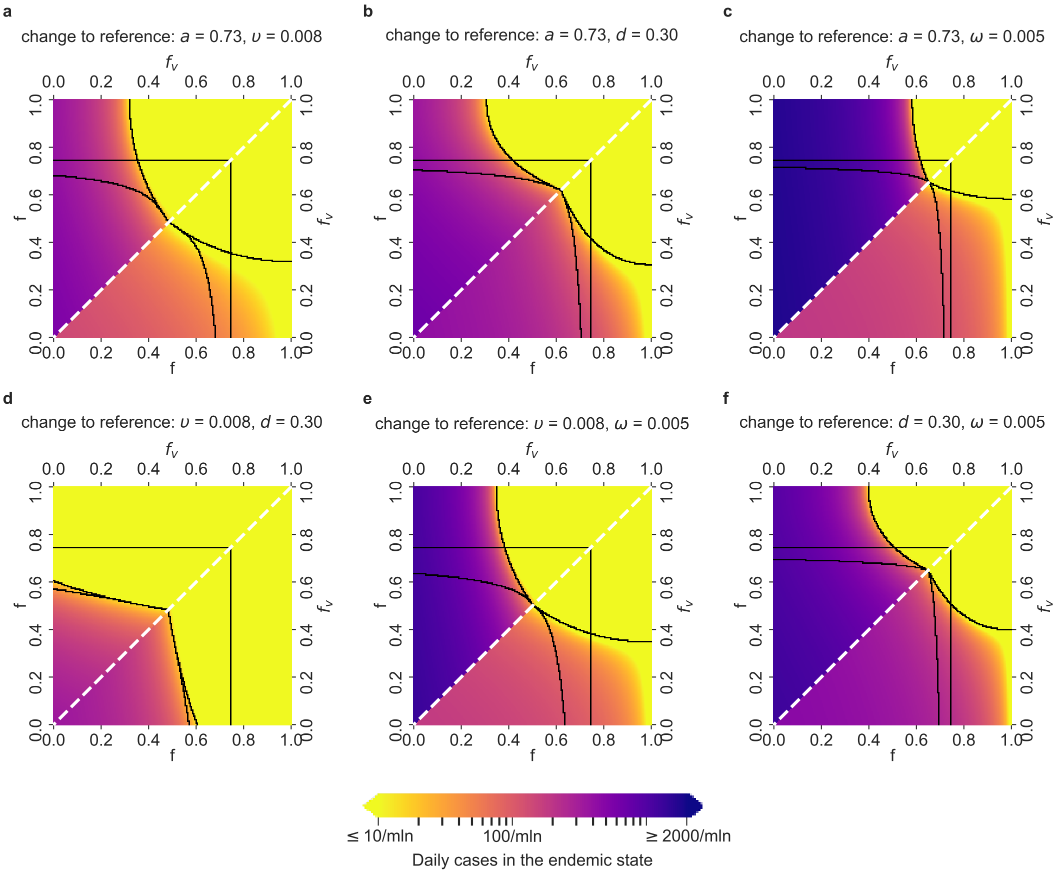


Supplementary Figure S7: **Daily COVID-19 infection cases in the endemic state, for parameter setups with two changes w.r.t. the reference setup, for the Alpha variant.** Lower triangles show the daily infection numbers in the unvaccinated, and upper triangles in the vaccinated population in the endemic state of the epidemics, for the relevant $f - f_\mathrm{v}$ parameter space, where $f_\mathrm{v} \leq f$. Parameter setups in panels **a.–f.** as in Supplementary Fig. S4.

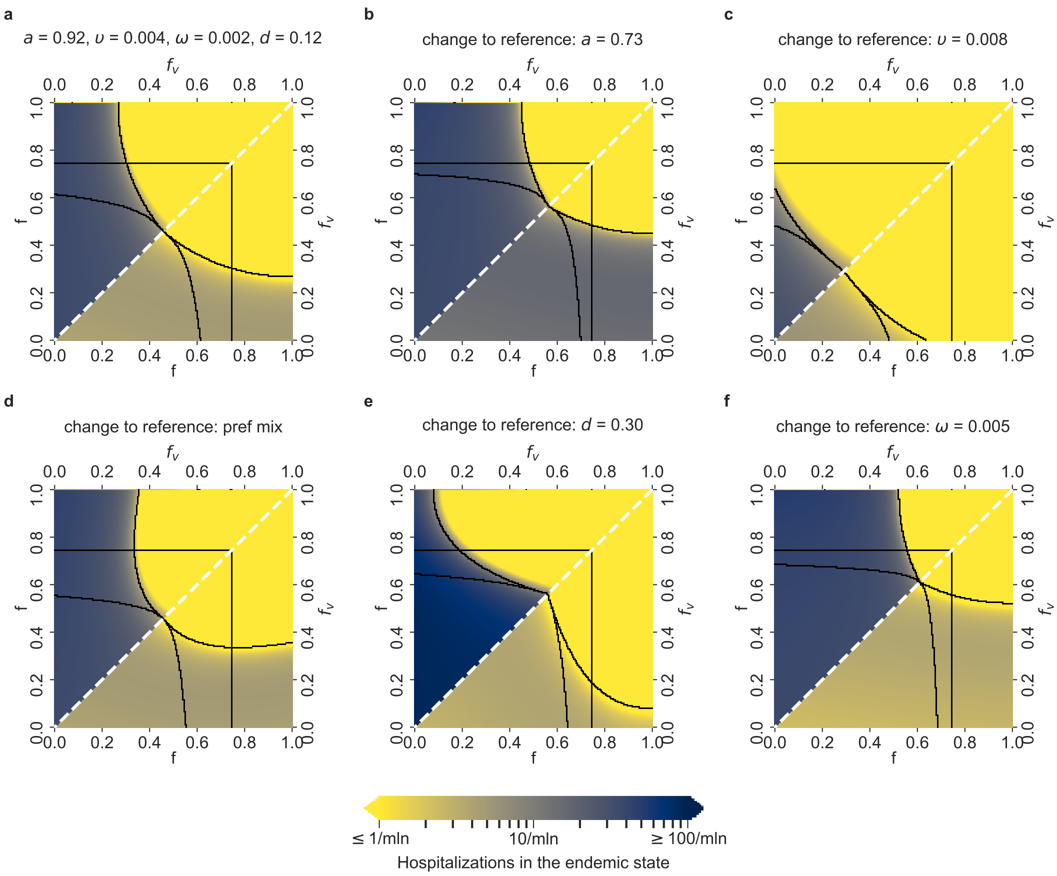


Supplementary Figure S8: **Daily COVID-19 hospitalized cases in the endemic state for different parameter setups and the Alpha variant.** Lower triangles show the daily hospitalized numbers in the unvaccinated population, and upper triangles in the vaccinated population, in the endemic state of the epidemic, for the relevant $f - f_v$ parameter space, where $f_v \leq f$. Parameter setups as well as the black borders that delimit the five regions are defined as in Supplementary Figure S2.

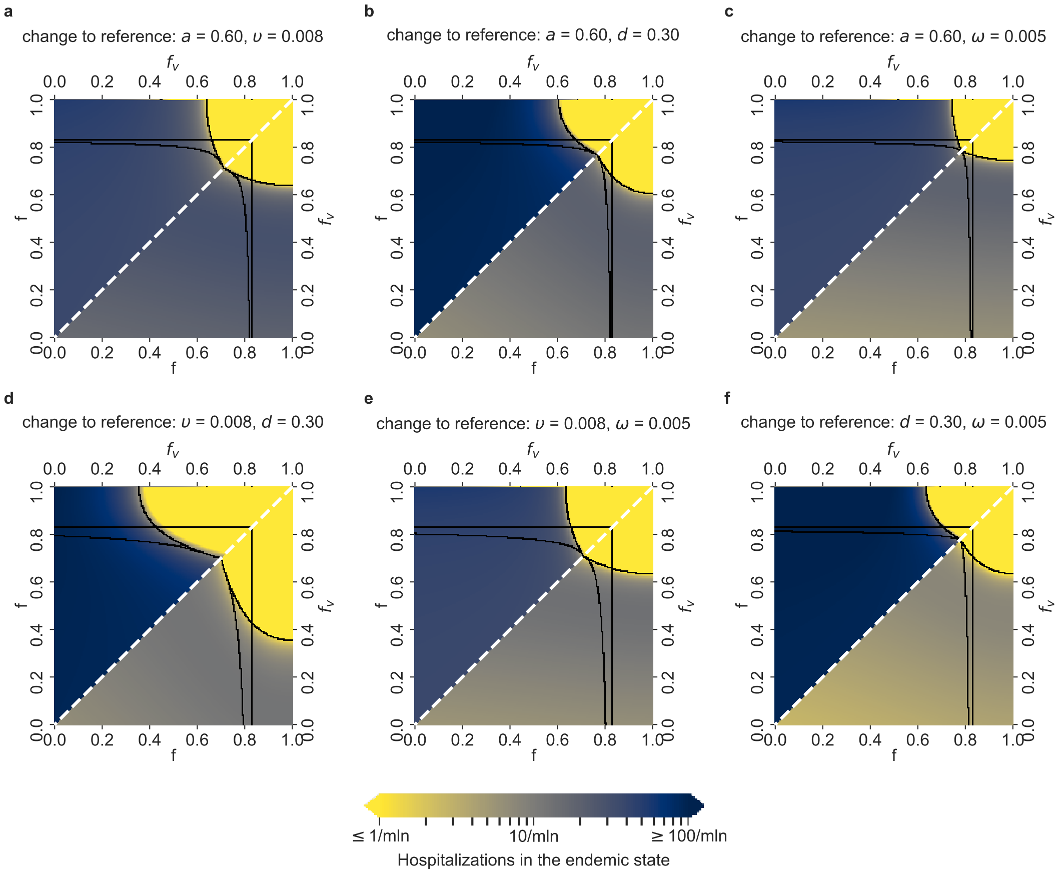


Supplementary Figure S9: **Daily COVID-19 hospitalized cases in the endemic state for parameter setups with two changes w.r.t. the reference setup, and the Delta variant.** Lower triangles show the daily hospitalized numbers in the unvaccinated population, and upper triangles in the vaccinated population, in the endemic state of the epidemic, for the relevant $f - f_v$ parameter space, where $f_v \leq f$. Parameter setups as well as the black borders that delimit the five regions are defined as in Supplementary Figure S3.

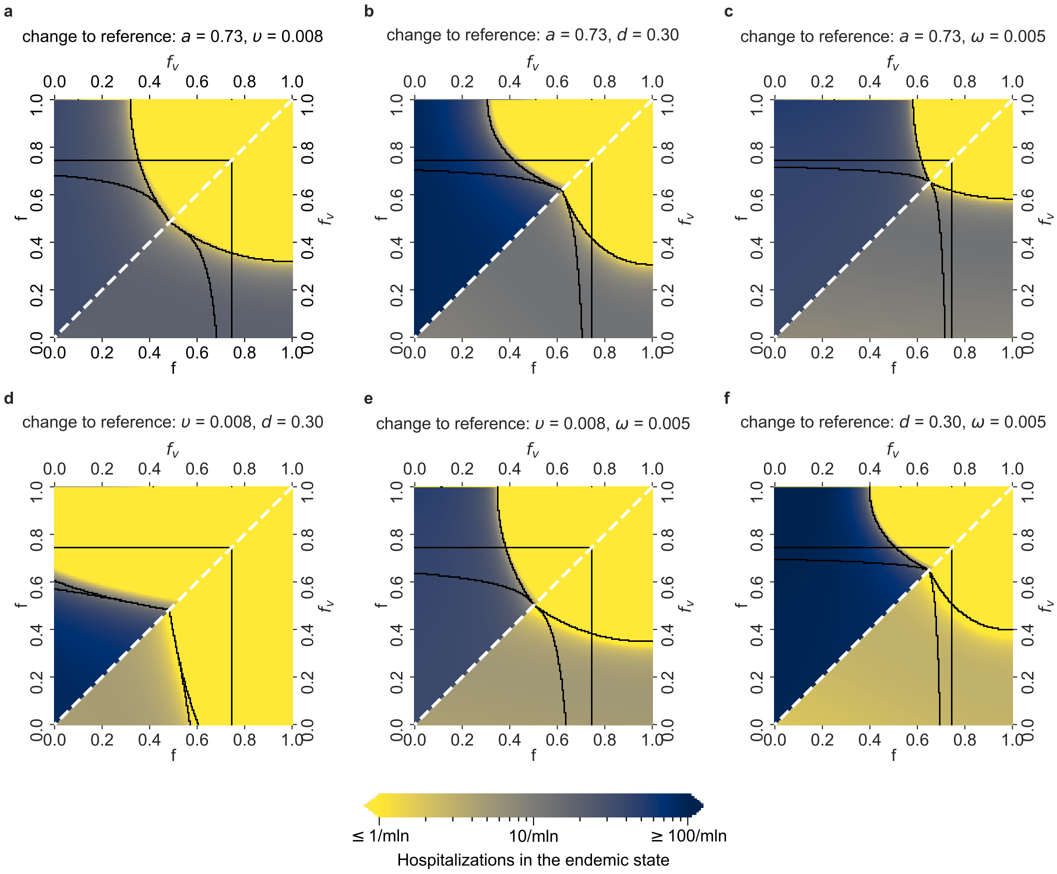


Supplementary Figure S10: **Daily COVID-19 hospitalized cases in the endemic state for parameter setups with two changes w.r.t. the reference setup, and the Alpha variant.** Lower triangles show the daily hospitalized numbers in the unvaccinated population, and upper triangles in the vaccinated population, in the endemic state of the epidemic, for the relevant $f - f_v$ parameter space, where $f_v \leq f$. Parameter setups as well as the black borders that delimit the five regions are defined as in Supplementary Figure S4.
